# European Survey on Citizens’ Attitudes towards Personalized Medicine, Genetic Testing, and Health Data Sharing: Design and Delivery

**DOI:** 10.1101/2024.02.02.24302142

**Authors:** Francesco Andrea Causio, Flavia Beccia, Loes Lindiwe Kreeftenberg, Giovanna Elisa Calabrò, Roberta Pastorino, Stefania Boccia, Carla van El

## Abstract

Personalized medicine leverages genetic, environmental, and lifestyle information to enhance the efficacy and safety of medical interventions. The shift from traditional one-size-fits-all approaches to a more individualized strategy signifies a major evolution in healthcare systems globally, promising improved patient outcomes and cost-effectiveness. A critical aspect of implementing personalized medicine is the role of public participation and their attitudes toward sharing health data. With growing concerns over data privacy, understanding and addressing public perceptions is essential. For this purpose, we designed and conducted a cross-sectional survey in Europe to gather public attitudes toward personalized medicine and health data sharing. The addressed issues range from knowledge and views to means of data sharing, e.g., via digital health instruments and electronic health care records. These insights are crucial for developing policies and practices that can help build trust and foster a secure environment for healthcare data sharing.

## Introduction

In an era marked by unprecedented advances in biomedical science and technology, personalized medicine has emerged as a transformative paradigm in healthcare, promising to revolutionize how we prevent, diagnose, and treat diseases. Unlike the one-size-fits-all approach of traditional medicine, personalized medicine better tailors medical interventions to the individual patient’s unique genetic, environmental, and lifestyle profiles. This profound shift from a population-based to an individualized approach can also significantly enhance the efficacy and safety of medical treatments, thereby offering new hope to patients and opportunities for the healthcare industry. (1)

Adopting personalized medicine represents a pivotal moment in the evolution of healthcare systems worldwide. With a growing body of evidence demonstrating its capacity to improve patient outcomes and reduce healthcare costs, healthcare providers, policymakers, and researchers increasingly recognize its importance. (2, 3) This paradigm shift has led to a burgeoning ecosystem of precision diagnostics, targeted therapies, and data-driven healthcare delivery systems. However, the successful implementation of personalized medicine critically hinges upon the active participation of citizens and positive attitudes toward sharing their health data. (4)

In light of the growing awareness and criticism of data privacy issues among the public, it is evident that trust plays a crucial role in the context of data sharing in personalized medicine. (5, 6) Therefore, surveying citizens’ attitudes towards personalized medicine and health data-sharing becomes even more crucial. By building on earlier research, such as the Your DNA Your Say (YDYS) project (4) and a survey targeting the Italian public (7), we addressed issues concerning the general public’s perceptions and attitudes toward genetic testing, as well as various means of health data sharing, including healthcare records and data-generating apps. The gathered comprehensive and up-to-date insights on public perceptions, concerns, and expectations regarding personalized medicine and health data sharing will be invaluable in shaping policies and practices that address these concerns and help foster a trustworthy healthcare data-sharing environment. The survey was conducted as part of the “European network staff eXchange for integrating precision health in the Health Care Systems” (ExACT) project, funded by the European Union’s Horizon 2020 research and innovation program, RISE Marie Curie Actions, aimed to provide an overview of European Union (EU)-wide attitudes on personalized medicine and health data sharing. Moreover, this survey is part of the activities of a project financed by the Italian Center for Disease Prevention and Control of the Ministry of Health, aimed at drafting an Italian Genomic Strategy and guaranteeing national support for the European initiative 1+Million Genomes (1+MG) and Beyond 1+MG (B1MG).

### Survey design

The survey design involved five researchers from Amsterdam UMC in the Netherlands and Università Cattolica del Sacro Cuore in Rome, Italy: RP, CVE, FAC, FB, and SB. All communications happened in English, and the first version of the survey was in English. The survey was conceptualized from scratch, building upon previous studies conducted in the two centers on related subjects, such as knowledge and attitudes regarding genetic testing and other examples retrieved in the literature. (7, 8) The survey was divided into four modules: A) “Knowledge and views,” B) “Data use and sharing,” C) “Governance,” and D) “Citizens’ needs.” Each section had short introductory sections informing the respondents before investigating their viewpoints on these aspects.

The final version was validated by experts in both institutions and submitted to the ethical review boards of Amsterdam UMC, which did not object to the study (reference 2022.0214), and Fondazione Policlinico Universitario Agostino Gemelli IRCCS in Rome, where it was approved with ID 5047.

### Survey composition and description

We designed a branching survey to ensure the optimal delivery of questions to the respondents. (9) The survey comprises 37 questions distributed across four main modules. The survey text is available in the Supplementary material.

Module A: “Knowledge and Views” consisted of six main questions that assess respondents’ baseline knowledge and perspectives on personalized medicine concepts, including various genetic testing applications. Additionally, this module features three branching sections for an in-depth exploration of respondents’ viewpoints based on their initial responses.

Module B: “Data Use and Sharing,” is further divided into three distinct sections to explore attitudes towards various forms of data sharing: Section 1, “Sharing Data from Health Care Dossiers and Records,” comprises two questions to uncover individuals’ attitudes and practices regarding sharing health data derived from their medical records; Section 2, “Donating Health Data to a Research Institute or Biobank,” includes seven questions, this section probes respondents about their willingness to donate health data to research institutions or biobanks, delving into motivations, concerns, and perspectives; Section 3, “Actively Contributing Health Data via Apps,” encompasses five questions centered around respondents’ experiences and inclinations to actively contribute their health data through mobile applications or similar platforms. It seeks to understand the factors driving or hindering such participation.

Module C, “Governance,” encompasses five questions to assess respondents’ opinions on personalized medicine’s regulatory and ethical aspects. Module D: “Citizens’ Needs,” examines how respondents perceive personalized medicine and their access to information.

### Survey translation

To ensure the best possible distribution relating to regions in Europe, the survey was translated into Dutch, Italian, French, Spanish, German, Polish, Hungarian, and Romanian. The translation was performed by either native language speakers in the issuing institutions or by professional translators hired by the authors.

### Survey distribution

Researchers agreed to contract the private company YouGov to distribute the survey on their platform. YouGov is a global public opinion and data company whose platform complies with the highest standards for quality and research while ensuring participant privacy. YouGov’s methodology is compliant with GDPR standards and is detailed on their website (https://yougov.co.uk/about/panel-methodology). The survey distribution lasted approximately two weeks during April 2023. Respondents are invited to participate in a YouGov survey based on their demographic information. They can complete the survey on the YouGov platform using their credentials. There is no time limit for filling out the survey, and participants can pause and resume as needed. Upon completion, participants receive a small non-monetary token of appreciation as compensation.

### Limitations

Despite the efforts to design and deliver this survey in the best way possible, some limitations should be considered: the study was conducted online, potentially introducing a bias toward individuals comfortable with internet usage. The survey was translated into a restricted number of languages, excluding individuals from different countries or linguistic backgrounds. The survey was not designed to be representative of the entire European population, limiting the generalizability of findings to other regions or countries. Participants’ responses were self-reported, which could introduce inaccuracies or dishonesty.

## Conclusion

This paper outlines the development of our cross-sectional online survey, which assesses European citizens’ attitudes toward personalized medicine and health data sharing. Administered to the general public across eight EU countries, the survey provides insights for future policymaking. Given the pivotal role of personalized medicine and health data sharing in shaping healthcare’s future, this survey addresses knowledge gaps in the field. This project represents the most extensive study to date on public perspectives on genomic data sharing in the EU.

## Future perspectives

In the future, personalized medicine is poised to revolutionize healthcare by tailoring treatments to individual patients, enhancing effectiveness and well-being. As evidence of its benefits accumulates, we anticipate increased acceptance among healthcare providers and policymakers, further driving its integration into mainstream healthcare systems. These developments foster a dynamic ecosystem of precision diagnostics, therapies, and data-driven healthcare solutions. The critical elements of building and maintaining trust in data sharing must be emphasized to help ensure its success, including transparency and robust privacy protection. Ongoing research and policy development will be pivotal in shaping the path forward.

## Data Availability

All data produced in the present study are available upon reasonable request to the authors

## Acknowledgments

We thank the respondents to the survey who took the time to answer the questions on personalized medicine. We thank the Italian Ministry of Health for supporting the activities of the National Center for Disease Prevention and Control (CCM) projects related to genomics and omics sciences. We also thank the numerous specialists involved in the CCM2021 project who are working daily in Italy for the implementation of omics sciences in the National Health System and for the health of our citizens.

