## Supplementary material for "European Survey on Citizens’ Attitudes towards Personalized Medicine, Genetic Testing, and Health Data Sharing: Design and Delivery"

Intro

The following survey is on the topic of personalised medicine/b. It has been developed as part of the ExACT project (“European network staff eXchange for integrAting precision health in the health Care sysTems” ), funded by the European Commission under the Horizon 2020 Programme. The consortium consists of academic partners, such as university medical centers, and non-academic partners, such as the European Public Health Association EUPHA. For more information, see
Partners (exactproject.net).

The survey has been distributed in collaboration with ExACT partners and the Active Citizenship Network.
Your YouGov account will be credited with points after completing the survey. Please remember that all answers are anonymous. The results will be stored for 15 years and analysed in accordance with the European Data Protection Regulation. We will take all necessary steps to preserve your anonymity during every stage of the research. More information is available in our privacy and cookies notice.
The survey should take around 15 minutes minutes to complete. You can restart from the point you stopped at any moment.
Please click here to continue.

Privacy notes
IT: https://it.yougov.com/about/privacy/
ES: https://es.yougov.com/about/privacy/
FR: https://fr.yougov.com/find-solutions/a-propos/politique-de-confidentialite/
DE: https://yougov.de/unternehmen/terms-combined/#/agb
NL: https://account.yougov.com/nl-nl/account/privacy-policy
HU: https://account.yougov.com/hu-hu/account/privacy-policy
PL: https://account.yougov.com/pl-pl/account/privacy-policy
RO: https://account.yougov.com/ro-ro/account/privacy-policy

**Module A: Knowledge and views**

Many people become ill at some point in their lives. For some disorders, our genes (i.e genetic predisposition) can increase the risk of becoming ill. However, also lifestyle and environment play a role – such as addictions, diet, or physical activity.
Tailoring prevention and care to each individual is the main aim of personalized medicine. The aim is to improve health or avoid chronic diseases by using information on individuals' genes and lifestyle. To better understand disease mechanisms, researchers need to collect a lot of information on the genes and health of many people. The results could be used to directly orientate medical care (primary use of data) or it could be used in research to gather more knowledge for future medical actions (secondary use of data).
In this survey we would like to know about your views/b about Personalized medicine; genetic testing; data sharing; governance of the research and introduction in health care. Remember there are no right or wrong answers, and your responses will remain anonymous.

It is now possible to entirely analyse a person's genome, or genetic material. This has the potential to provide detailed information about individual health. Researchers hope that this can be used to guide individuals to better manage their health, both preventing diseases and choosing the best medical care for them.
Click "" to continue.

1) Which of the following had you ever heard about, prior to this survey?
Please select one answer per row.

| | Yes, I had heard of this | No, I had never heard of this | Don't know |
|---|---|---|---|
| Personalized medicine | | | |
| Big data | | | |
| Genetic testing | | | |

*Only to those who answered "Yes, I had heard of this" about personalized medicine:*
1a) You said you had heard about personalized medicine before. How did you hear about it?
- o I or someone I know have a medical condition
- o My work is related to healthcare (i.e. policymakers, healthcare workers, …)
- o I have come to learn about this topic via other means
- o Don't know

*Only to those who answered "Yes, I had heard of this" about genetic testing:*
1b) You said you had heard about genetic testing before. How did you hear about it?
Please select all that apply.
- o From my school or studies
- o From my work as researcher
- o From my work related to health care
- o From my work, unrelated to research or healthcare
- o I or someone I know has a hereditary disorder
- o I or someone I know has had genetic testing to inform treatment for a disease

- I know about genetic testing to learn about ancestry, fitness or diet
- Other
- Don't know / Can't remember

*Only to those who answered "Yes, I had heard of this" about personalized medicine AND genetic testing:*

2) Were you aware about the connection between genetic testing and personalized medicine/b prior to this survey?
- Yes
- No
- Don't know / not sure

3) Which of the following uses of genetic testing for medical purposes have you ever heard about prior to this survey?
Please select all that apply.
- Diagnosis of a disease
- Assessment of risk to develop a specific disease, for the disease to be prevented or treated at an early stage
- Choice of the treatment for a disease (e.g. the choice of chemotherapy in case of cancer)
- Assessment of a specific drug in a person (e.g. to avoid wrong dosage or adverse effects)
- Assessment of the risk of passing a genetic disease to future children
- Prenatal testing during pregnancy
- Heel prick of newborn baby to detect a genetic disorder
- Other
- None

4) If a genetic test were available at low cost, for what purposes you would support the test being offered by the health care system of your country, and would you consider having such testing?
Please select one answer per row.

| | I would support the test being made available by the health care system of my country, and I would consider taking such test | I would support being made available by the health care system of my country, but I would NOT consider taking such test | I do not support the test being made available in by the health care system of my country | Don't know |
|---|---|---|---|---|
| To diagnose a serious genetic disease | | | | |
| To assess the predisposition or risk for you to develop a | | | | |

| |
|---|
| specific disease in the future |
| To choose the most effective treatment or a treatment with the lowest risk of potential adverse effects |
| Before pregnancy, to assess the risk of future parents transmitting a predisposition for a specific disease to their future children |
| During pregnancy, to diagnose or assess the risk of a serious disease in the foetus? |

*Only to those who answered, "I do not support the test being made available in by the health care system of my country" on any option in question 4:*
4a) You have said you would oppose genetic tests being made available for the following purposes. Could you please let us know why?

- o To diagnose a serious genetic disease
- o To assess the predisposition or risk for you to develop a specific disease in the future
- o To choose the most effective treatment or a treatment with the lowest risk of potential adverse effects
- o Before pregnancy, to assess the risk of future parents transmitting a predisposition for a specific disease to their future children
- o During pregnancy, to diagnose or assess the risk of a serious disease in the foetus?

5) In which cases would you like to know the results of a genetic test you have undergone?

- o Always
- o Only if preventive measures or treatments are available from the results
- o I would not want to know myself, but I would want my doctor to know
- o I would not want myself or others to know
- o Don't know

6) How do you think that genetic testing for disease could impact individuals?
Please select one option per row.

| | Definitely agree | Somewhat agree | Neither agree nor disagree | Somewhat disagree | Definitely disagree | Don’t know |
|---|---|---|---|---|---|---|
| People would worry more about their health | | | | | | |
| People would be more informed about their health | | | | | | |
| People would be more empowered to make health-related decisions | | | | | | |
| People would feel more pressure to make certain lifestyle choices | | | | | | |
| People would feel more controlled by the government | | | | | | |

**Module B: Data Use and Sharing**

Data sharing is a contentious issue. On the one hand, it can raise concerns related to citizens' privacy, and it requires specific infrastructure and policies. On the other hand, it can be crucial to understand the mechanisms underlying a disease, and allow for a more personalised approach in medicine.
There are several ways people can share their data:

1. Allowing data from their health care dossiers to be shared with health care professionals to help them diagnose other patients (use in patient care) or with researchers (use for research);
2. Donating health data to a research institute or so-called biobank at their own initiative;
3. Actively sharing data from health apps they use.

In this section, we are interested in your views on these 3 forms of use and sharing of health-related data.

*1. Sharing data from health care dossiers and records*
In order to facilitate access of medical information, general practitioners and hospitals have medical health records of their patients, be it physical or electronic health records (EHR). Health portals are instruments created by governments to engage citizens and give them access to their health data via a secure online website. Using a secure username and password, patients can view health information such as recent doctor visits, discharge summaries, medications and lab results among others.

7) Were you aware of the existence of health/patient portals prior to this survey?

- o Yes
- o No
- o Don’t know

8) If people were given an option to share their data, would you be willing to share your data - including from any genetic tests - from your health care record or portal to benefit other patients or for medical research purposes?

- o Yes, to help health care professionals interpret findings and diagnose other patients
- o Yes, but only to support research into a disease I or members in my family suffer from
- o I would share health data with researchers, but not genetic data
- o No, I wouldn’t
- o Don’t know

*Only to those who answered, “Yes, to help health care professionals interpret findings and diagnose other patients”, “Yes, but only to support research into a disease I or members in my family suffer from” or “I would share health data with researchers, but not genetic data” in question 8:*
8a) You’ve said you would share health-related data. If you were given an option to share data from your healthcare systems’ records to research institutes, what type of institutes would you consider sharing your data with?

- o Only public ones (e.g. universities or research institutes from medical centers)

- Only private ones (e.g. pharmaceutical companies, private research institutes or other companies)
- Both public and private ones
- Don't know

*Only to those who answered, "No, I wouldn't" in question 8:*
8b) You've said you would not share health data from your records or portal. What are the reasons?
Please select all that apply

- I don't trust that my privacy will be respected when handling my data
- I don't trust my data will be used for private profit rather than to help society
- I am scared by what scientists may do with my genetic data
- I think I should be compensated (paid) for providing my data
- Other reasons (please specify)
- Don't know

*2. Donating health data to a research institute or so-called biobank at one's own initiative*
People can donate samples (such as tissue from operation or blood draws) to be stored in so-called 'biobanks'. These samples can benefit research, but some have raised privacy concerns. To safeguard privacy, medical data are currently stored and used without scientists knowing the names and personal details of the patient or donor of the data. But, if people that contribute to the research want to know their individual results themselves, there could be an option to reconnect the original record to the results – a third party that can be trusted and that has no connection to the research can be asked to retrieve the donor's results for them.

9) Would you share your personal genomic data with biobanks or research institutes?

- Yes, for the sake of contributing to science
- Only provided some reassurance or information are granted to me
- Only provided I would be adequately compensated monetarily
- No, in any case I wouldn't
- Don't know

*Only to those who answered, "*Yes, for the sake of contributing to science," *"*Only provided some reassurance or information are granted to me," or "Only provided I would be adequately compensated monetarily" *in question 9:*
10) You have said that, under some circumstances, you would share your personal genomic data with biobanks. If you did, what kind of information on the results would you like to receive, if that would be available/possible?

- I would NOT want to receive any feedback on my results
- I would donate for research on a specific disease and would like to receive information on that disease only
- I would want relevant findings on my genes to be reported to me
- I would want relevant findings regarding my health or lifestyle reported to me
- I would like to know about ANY individual results
- Don't know

*Only to those who answered "*Yes, for the sake of contributing to science," *"*Only provided some reassurance or information are granted to me," or "Only provided I would be adequately compensated monetarily" *in question 9:*

11) Which types of research institutions (i.e biobanks) would you share your data with?

- Only public ones (e.g. universities or research institutes from medical centers)
- Only private ones (e.g. pharmaceutical companies, private research institutes or other companies)
- Both public and private ones
- know

12) Do you agree or disagree that the privacy of individuals participating in genomic and other health care studies is adequately protected in your country?

- Definitely agree
- Somewhat agree
- Neither agree nor disagree
- Somewhat disagree
- Definitely disagree
- Don't know

13) In modern science, biobanks often collaborate with universities and hospitals to share health-related information and do research.
Provided privacy policies prevent you from being identified, would you agree that your anonymized data can be shared with other countries?

- Yes, I would support such data sharing with other countries even outside the EU, if that were overseen by ad-hoc regulations
- Yes, I would support such data sharing in the if that were overseen by ad-hoc regulations, but only within the EU
- No, I would not agree
- Don't know

14) In the EU, collaboration between universities and private companies is stimulated to boost innovation and economic growth. Private companies may further develop findings from research in academia, for instance to market a drug or test.
Would you agree with the use of your anonymized personal genetic and lifestyle data for such commercial purposes?

- Yes
- No
- Don't know

15) Which, if any, of the following do you perceive as potential risks related to sharing your health data with a biobank?
Please select all that apply.

- Data being hacked or used for unauthorised purposes
- Data being wrongfully reported
- Being identified from my data

- Identity theft
- Data being used by the government for discrimination
- Health-related stigma, which has negative consequences in the workplace, in obtaining insurance, or in society
- Data being used for commercial gain
- My genetic information being linked to a crime committed by me or someone else
- Other
- Not applicable: I do not perceive any risks from this
- Don’t know

Intro

*3. Actively contributing your health data via apps*

Mobile e-health apps are software programs that run on for instance smartphones and other mobile communication devices. Consumers can use these apps to manage their own health and wellness, such as to monitor their caloric intake or sugar levels. Other apps aim to help health care professionals improve and facilitate patient care.

16) Would you be interested in using personal health care apps monitoring your health (e.g. heart rate, exercise, response to medications)?

- Yes, I already use such apps
- Yes, I might use such apps in the future
- No, I am not interested
- Don’t know

*Only to those who answered* “Yes, I already use such apps” or “Yes, I might use such apps in the future” to question 16

17) What information would you like to be provided to you before using these health apps? Please select all that apply.

- Clear description of the purposes
- Information about the developers
- Information about data usage and who can access the data
- Information on rights to delete my data
- Information on data portability (use on other devices or platforms)
- Information on the storage of data
- Privacy policies
- Other
- Don’t know

18) If it were possible in your country to share the data from your personal health apps with your medical record/health portal so your health care provider or doctor would have access, would you be willing to share your data?

- Yes
- No
- Don’t know

19) And would you be willing to share the data from your personal health apps with research institutes or biobanks?

- Yes, but only with public ones
- Yes, but only private ones
- Both public and private ones
- No
- Don’t know

20) Which, if any, of the following do you perceive as potential risks related to sharing your data in health apps?
Please select all that apply.

- Data being hacked or used for unauthorised purposes
- Data being wrongfully reported
- Being identified from my data
- Identity theft
- Data being used by the government for discrimination
- Health-related stigma, which has negative consequences in the workplace, in obtaining insurance, or in society
- Data being used for commercial gain
- My genetic information being linked to a crime committed by me or someone else
- Other
- Not applicable: I do not perceive any risks from this
- Don’t know

**Module C: Governance**

Thanks for your responses so far. In this section, we are interested in your views on how personalized medicine should be controlled.
Citizens' and patients' involvement is key to policy development. Making patients aware of possible healthcare opportunities requires them to know more about healthcare data, including genetic data, biobanks and how they are governed. Good governance implies transparency and accountability, besides implementation of oversight mechanisms.

21) In your opinion, who should be involved in making policies and regulations in the field of personalized medicine?
Please select all that apply.

- o Policymakers
- o Citizens and patients, directly or via their representatives (e.g. associations, groups…)
- o Healthcare professionals
- o Religious or spiritual entities
- o Other
- o Don't know

22) Careful handling of health data is key. In EU countries, the General Data Protection Regulation (GDPR) recognises data concerning health as a special category of data.
Which of the following frameworks regulation concerning health data and personalized medicine you aware of?

| | I am aware of its provisions on health data | I have heard of it, but I am not aware of its provisions on health data | I have never heard of it | Don't know |
|---|---|---|---|---|
| GDPR | | | | |
| National regulation of my country | | | | |

23) Biobanks are currently overseen by committees consisting of representatives of scientists, patients, and ethicists who decide what type of research to conduct and with what institutions they share their data.
Do you think biobanks should be additionally overseen by dedicated national or supranational institutions?

- o Yes
- o No
- o Don't know

*Only to those who answered* "Yes" to question 23:
24) You have said you think biobanks should be additionally overseen by dedicated national or supranational institutions. At which level do you think these institutions should be?

- o At global level
- o At European Union level
- o At national level
- o Don't know

25) Do you think citizens using health apps should have a say about the use of their data for research purposes?

- Yes, I think one should be asked permission for every request for data use
- Yes, I think a general consent to use data for specified research purposes would be enough
- No
- Don’t know

**Module D: Citizens' needs**

Finally, a few questions on how to inform citizens regarding personalized medicine.

26) How reliable would you say each of the following media is to deliver information about personalized medicine?
Please select one answer per row.

| | Very reliable | Somewhat reliable | Not very reliable | Not reliable at all | Don't know |
|---|---|---|---|---|---|
| TV | | | | | |
| Newspapers, magazines, broadsheets | | | | | |
| Specialized websites | | | | | |
| Social media posts | | | | | |
| Radio broadcasts | | | | | |
| Schools and university | | | | | |
| Institutional communication | | | | | |
| Healthcare providers (doctors, hospitals…) | | | | | |

27) Do you think information about personalized medicine is easily accessible by citizens?
- Definitely
- Somewhat
- Not really
- Not at all
- Don't know

28) Do you think you have adequate knowledge about personalized medicine?
- Definitely
- Somewhat
- Not really
- Not at all
- Don't know

*Only to those who answered "Not really" or "Not at all" to question 28:*
29) You have said you do not feel like you have adequate knowledge about personalized medicine. What do you feel you would need to know more about?
- Data management and privacy
- Research benefits from personalized medicine
- Clinical outcomes from application of personalized medicine
- How personalized medicine can be useful to me
- How personalized medicine can help reduce the cost of healthcare for taxpayers
- Other (specify)
- Don't know

30) Do you think that personalized medicine would be beneficial for individuals' health?

- o Definitely
- o Somewhat
- o Not really
- o Not at all
- o Don't know

31) In general terms, are you in favour or against the application of personalized medicine into clinical practice?

- o Strongly in favour
- o Somewhat in favour
- o Neutral
- o Somewhat against
- o Strongly against
- o Don't know

32) Are there any additional topics regarding personalised medicine that you feel like should have been covered in this survey?

- o Yes (specify)
- o No
